# Serologic biomarkers and scoring systems for hepatic fibrosis in obese children with nonalcoholic steatohepatitis

**DOI:** 10.1101/2023.08.25.23294634

**Authors:** Jung Yeon Joo, In Hyuk Yoo, Hye Ran Yang

## Abstract

**Objectives:** The prevalence of nonalcoholic steatohepatitis (NASH) is increasing with the increasing prevalence of childhood obesity. Although NASH has a high risk of progression to liver fibrosis and cirrhosis, few studies have reported on noninvasive markers for predicting hepatic fibrosis in children. This study aimed to evaluate and compare the diagnostic accuracy of serologic biomarkers and scoring systems for hepatic fibrosis in obese children with NASH.

**Methods:** A total of 99 children were diagnosed with NASH based on liver biopsy findings, and divided into two groups according to the degree of liver fibrosis: mild (stage 0-1) or advanced (stage 2-4). Clinical and laboratory parameters and serum levels of hyaluronic acid and type IV collagen were measured. Moreover, the aspartate aminotransferase/platelet ratio index (APRI) and fibrosis-4 (FIB-4) score were calculated.

**Results:** Among the noninvasive markers, only serum type IV collagen level and FIB-4 revealed significant differences between the two fibrosis groups. The area under the receiver operating curve of each biomarker and scoring system was 0.80 (95% confidence interval [CI]: 0.70-0.90) for type IV collagen at an optimal cutoff of 148 ng/mL (sensitivity 69.8%, specificity 84.6%), followed by 0.69 (95% CI: 0.57-0.83) for APRI, 0.68 (95% CI: 0.56-0.80) for FIB-4, and 0.65 (95% CI: 0.53-0.77) for hyaluronic acid.

**Conclusions:** Type IV collagen as a single noninvasive serologic biomarker for hepatic fibrosis and FIB-4 as a hepatic fibrosis score are beneficial in predicting advanced hepatic fibrosis, and in determining proper diagnosis and treatment strategies before fibrosis progresses in obese children with NASH.

**Why was this study done?:** With the increase of the obese population of children and adolescents, the prevalence of nonalcoholic fatty liver disease is increasing. Non-alcoholic steatohepatitis has a high risk of developing liver fibrosis and cirrhosis. However, a single non-invasive serological marker that can predict fatty liver fibrosis in children and adolescents has not been clearly presented.

**What did the researchers do and find?:** We evaluate diagnostic accuracy of serological biomarkers and scoring systems for hepatic fibrosis in obese children with NASH.

**What do these findings mean?:** Type IV collagen is a useful single noninvasive serological biomarker for hepatic fibrosis. Therefore, it is useful in making appropriate diagnosis and treatment decisions before fibrosis progresses in obese children with non-alcoholic steatohepatitis.

## Background

The prevalence of obesity among children and adolescents worldwide, including South Korea, has increased in recent years [1]. Accordingly, the prevalence of obesity-related complications, such as nonalcoholic fatty liver disease (NAFLD) and metabolic syndromes is also rapidly increasing [2, 3].

NAFLD is a spectrum of diseases including simple steatosis, nonalcoholic steatohepatitis (NASH), liver fibrosis, and cirrhosis. NASH has a high risk of progression to advanced liver diseases such as liver fibrosis and cirrhosis, even in children [4–6].

Liver fibrosis is a dynamic process involving the two conflicting processes of fibrogenesis and fibrolysis. These processes result in the deposition of collagen and extracellular matrix proteins in tissues [7]. Therefore, the degree of liver fibrosis is an important prognostic factor for determining the prognosis and timing of chronic liver disease.

Liver biopsy is the gold standard method for the diagnosis of hepatic fibrosis [8, 9]. However, it has limitations such as complications due to invasive techniques, sample errors and cost. Therefore, the research interest on noninvasive diagnostic methods is increasing. However, studies on noninvasive serologic markers for liver fibrosis screening in NASH have mainly been conducted in adult patients [10] and there are few studies in children and adolescents. Furthermore, no single biochemical marker for liver fibrosis has been reported, and studies on noninvasive fibrosis scoring systems are still insufficient.

Therefore, the aim of this study was to evaluate and compare the diagnostic accuracy of serologic markers related to the pathophysiology of liver fibrosis and scoring systems for hepatic fibrosis, according to the severity of liver fibrosis in obese children with NASH.

## Methods

### Ethics statements

This retrospective study was approved by the institutional review board of Seoul National University Bundang Hospital (approval no. B-1909-562-104). The requirement for individual patient consent was waived as this study was based on anonymized data from electronic medical record (EMR).

### Study population and data collection

This study was a retrospective review of the medical records of 96 obese children and adolescents aged < 18 years with biopsy-proven NASH, who visited the Department of Pediatric Gastroenterology and Hepatology at Seoul National University Bundang Hospital between July 2003 and May 2019.

We planned the study in August 2019 and conducted the analysis between September and December 2019.

The medical data of each study subject, including age, sex, weight, height, body mass index (BMI), abdominal circumference (AC), laboratory test values, radiologic findings, and histopathologic findings, were reviewed and retrospectively analyzed.

Obesity was defined as a BMI higher than the 95th percentile for age and sex. BMI was calculated as weight (kg) divided by the height squared (m^2^). BMI was determined according to age and sex based on the 2017 Korean National Growth Chart [11].

All study subjects were diagnosed with NAFLD after excluding other causes of chronic hepatitis, such as hepatitis (A, B, C, E) virus, cytomegalovirus, Epstein-Barr virus, Wilson’s disease, metabolic disease, autoimmune hepatitis, and drug toxicity.

The study subjects were divided into two groups according to the histopathologic grading and staging of NAFLD. Hepatic fibrosis of stage 0-1 was defined as mild fibrosis, and fibrosis of stage 2-4 was defined as advanced fibrosis.

### Laboratory tests and serologic markers of hepatic fibrosis

All study subjects underwent laboratory tests including fasting glucose, insulin, hemoglobin A1c, total cholesterol, triglyceride, high-density lipoprotein cholesterol, low-density lipoprotein cholesterol, and apoprotein A1 and B levels. Serum aspartate aminotransferase (AST), alanine aminotransferase (ALT), alkaline phosphatase, γ-glutamyl transpeptidase, total bilirubin, albumin, and prothrombin time were also measured.

Among potential biomarkers based on the pathogenesis of hepatic fibrosis, type IV collagen and hyaluronic acid (HA) were additionally measured at the time of diagnosis in all study subjects. Type IV collagen was measured using a radioimmunology assay, and HA was measured using a latex agglutination immunoassay.

### Hepatic fibrosis scoring systems

The AST/ALT ratio was calculated as the ratio of AST to ALT [12]. The AST/platelet ratio index (APRI) was calculated as follows: (AST level / AST upper level of normal / platelet count) × 100 [13]. Fibrosis-4 (FIB-4) was calculated as (age × AST level / platelet count × √ALT) [14]. PGA index combines the measurement of the prothrombin index, GGT level, and apolipoprotein A1 level [15].

### Radiologic investigations of the liver

The presence of fatty liver was evaluated in each patient using abdominal sonography and/or noncontrast abdominal magnetic resonance imaging. On abdominal sonography, the degree of fatty liver was defined as mild, moderate, and severe.

### Histopathologic findings of the liver and staging of fibrosis

All study subjects underwent percutaneous needle liver biopsy under local anesthesia. Specimens were evaluated to diagnose NAFLD and to assess the stage of liver fibrosis according to the Knodell scoring system [16, 17]. The stages of fibrosis were categorized into four groups: none (stage 0), perisinusoidal or periportal fibrosis (stage 1), perisinusoidal and portal/periportal fibrosis (stage 2), bridging fibrosis (stage 3), and cirrhosis (stage 4) [17].

### Statistical analysis

The results are expressed as mean ± standard deviation. The data were analyzed using PASW Statistics (version 22.0; SPSS Inc., Chicago, IL, USA). The Kruskal-Wallis test was used to compare three or more quantitative nonparametric variables. Chi-square tests were used to compare categorical variables. For all statistical analyses, a two-sided *p-*value of < 0.05 was considered statistically significant.

Receiver-operating characteristic (ROC) curves were used to evaluate optimal cutoff levels, and the area under the ROC curve (AUROC) was calculated.

## Results

### Patient characteristics

A total of 96 children diagnosed with NASH were included in the study. Among the 96 total patients, 78 (81.3%) were boys and 18 (18.7%) were girls, with a mean age of 12.4 ± 3.1 years. The mean AC was 91.5 ± 11.7 cm. The mean BMI was 27.0 ± 4.6 kg/m^2^. The clinical characteristics of obese children with NAFLD are shown in Table 1.

**Table 1.**
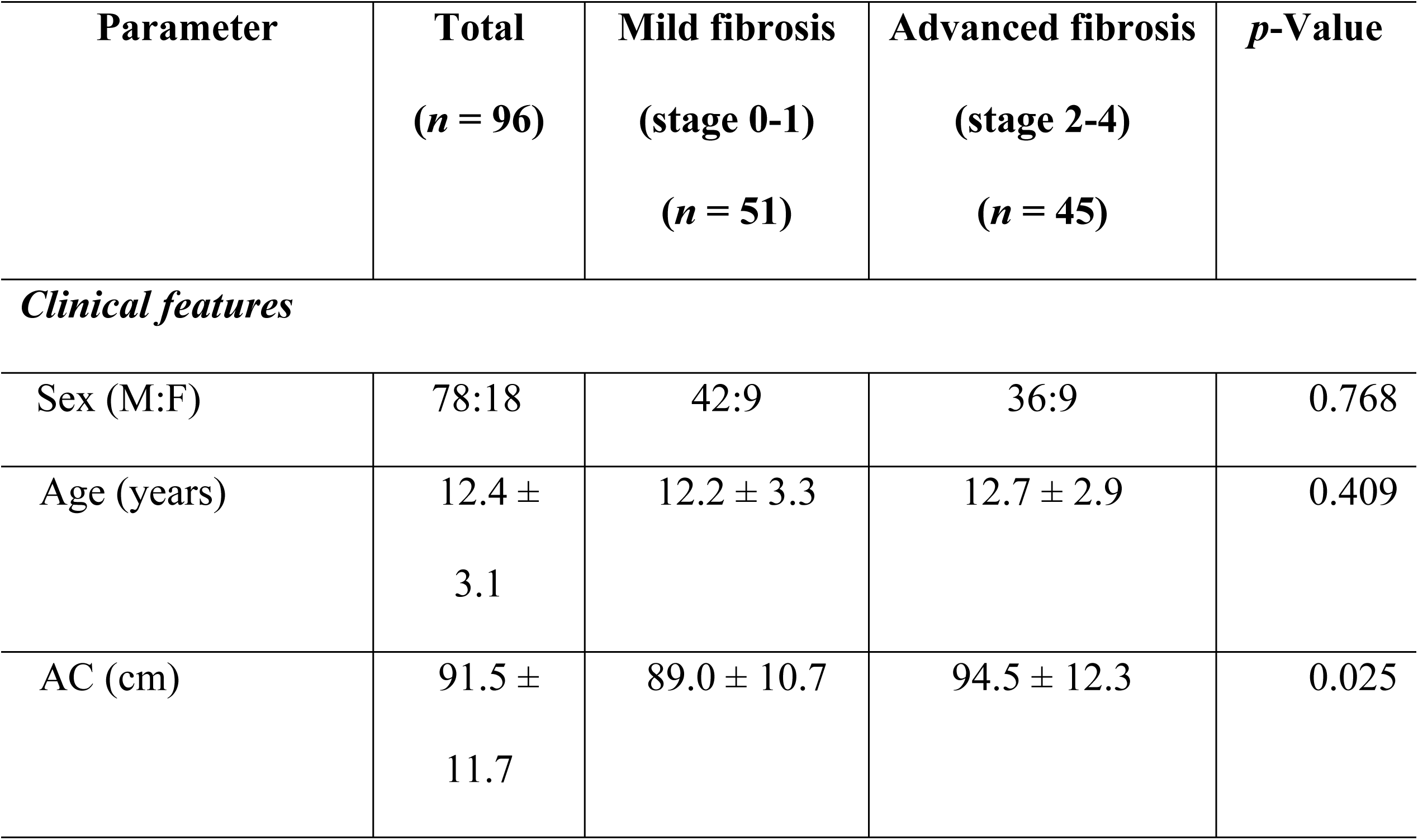
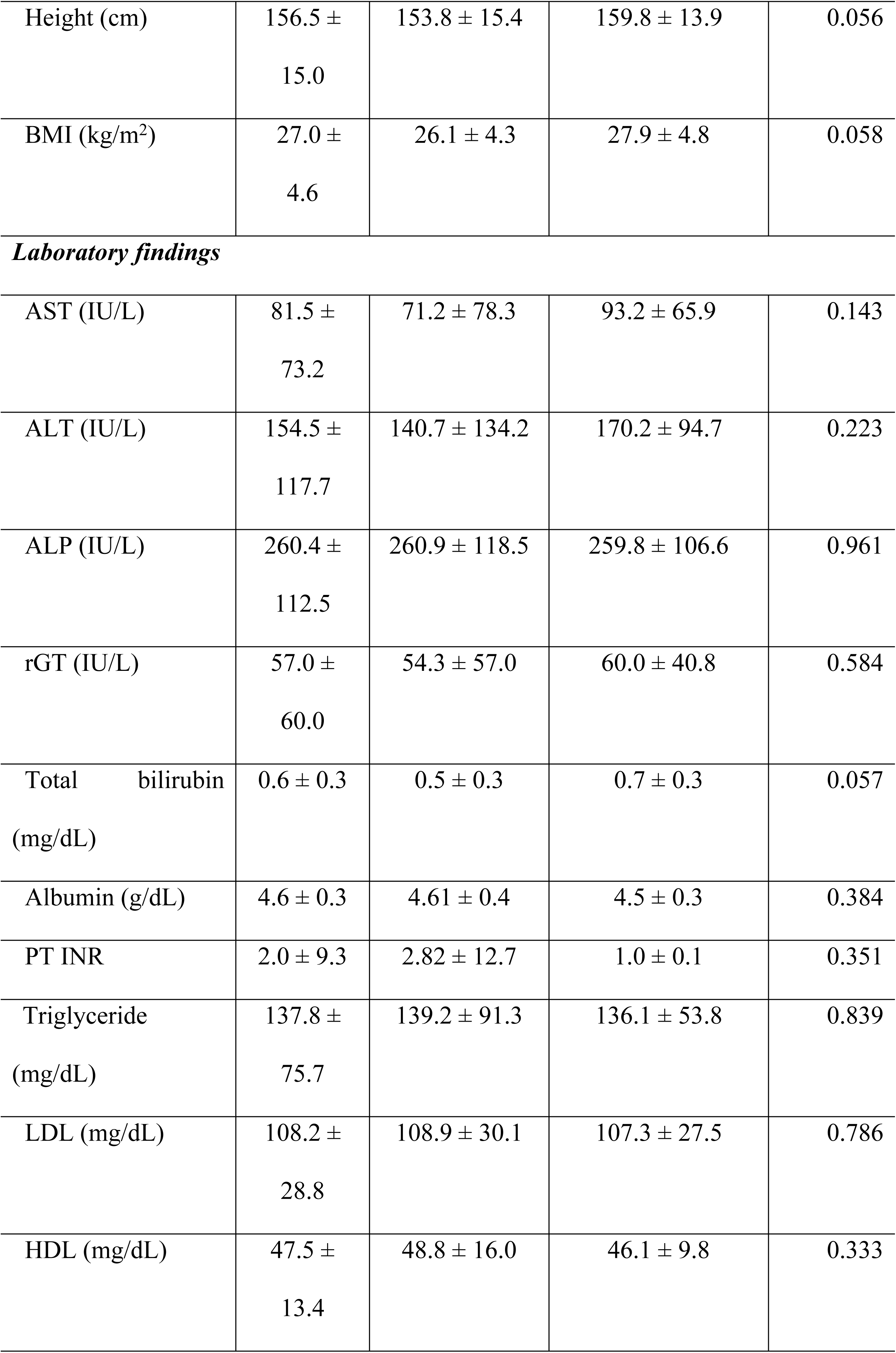
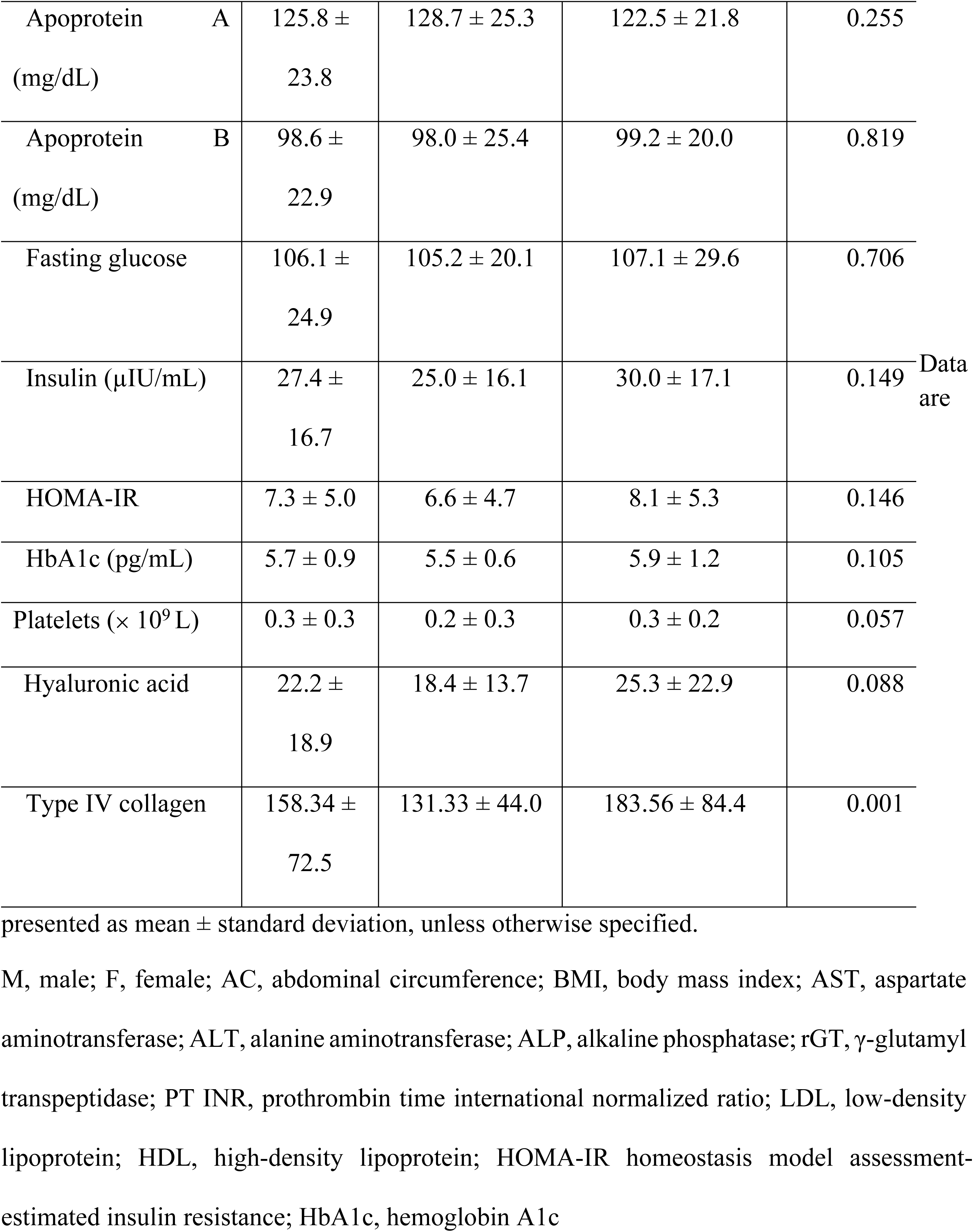
Comparison of clinical features and laboratory parameters between mild fibrosis (stage 0-1) and advanced fibrosis (stage 2-4) in obese children with nonalcoholic steatohepatitis.

### Comparison of clinical and laboratory parameters between mild and advanced fibrosis

Of the 96 patients diagnosed with NASH, 51 were in the mild fibrosis group and 45 were in the advanced fibrosis group. Significant differences were observed in AC measured at diagnosis between the mild fibrosis group and the advanced fibrosis group (*p* = 0.008) (Table 1). However, the other clinical and laboratory parameters did not significantly differ between the two groups (Table 1).

### Comparison of noninvasive hepatic fibrosis markers between mild and advanced fibrosis

When comparing the serum levels of biomarkers of hepatic fibrosis between the mild and advanced fibrosis groups, a statistically significant difference was found in type IV collagen (131.3 ± 44.0 vs. 183.6 ± 84.4 ng/mL, *p* = 0.001) (Table 2). The level of HA did not significantly differ between the two groups (*p* = 0.088).

**Table 2.**
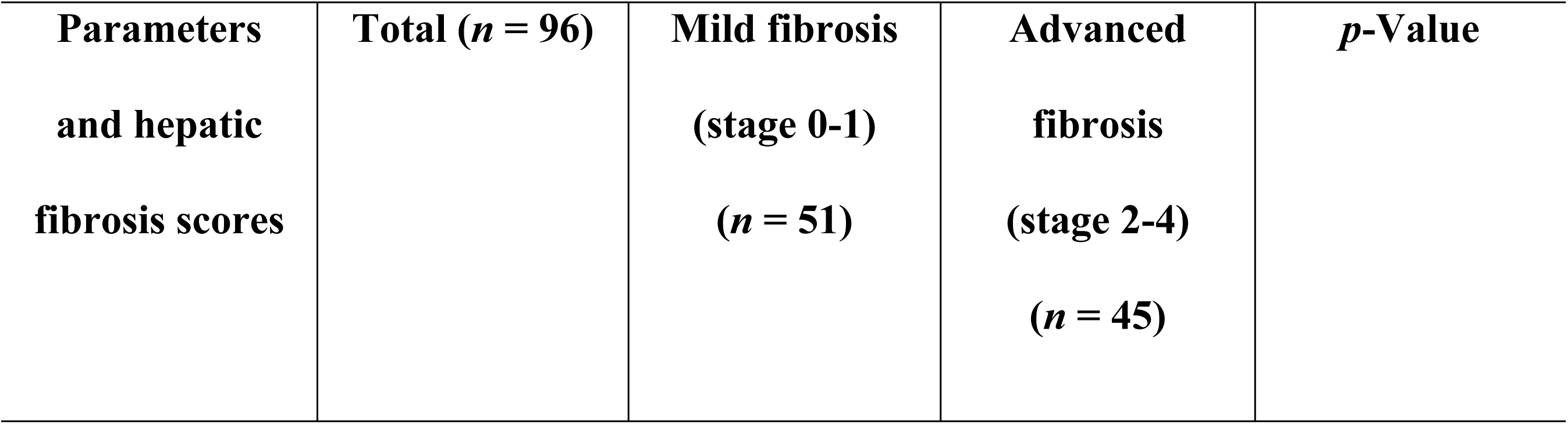
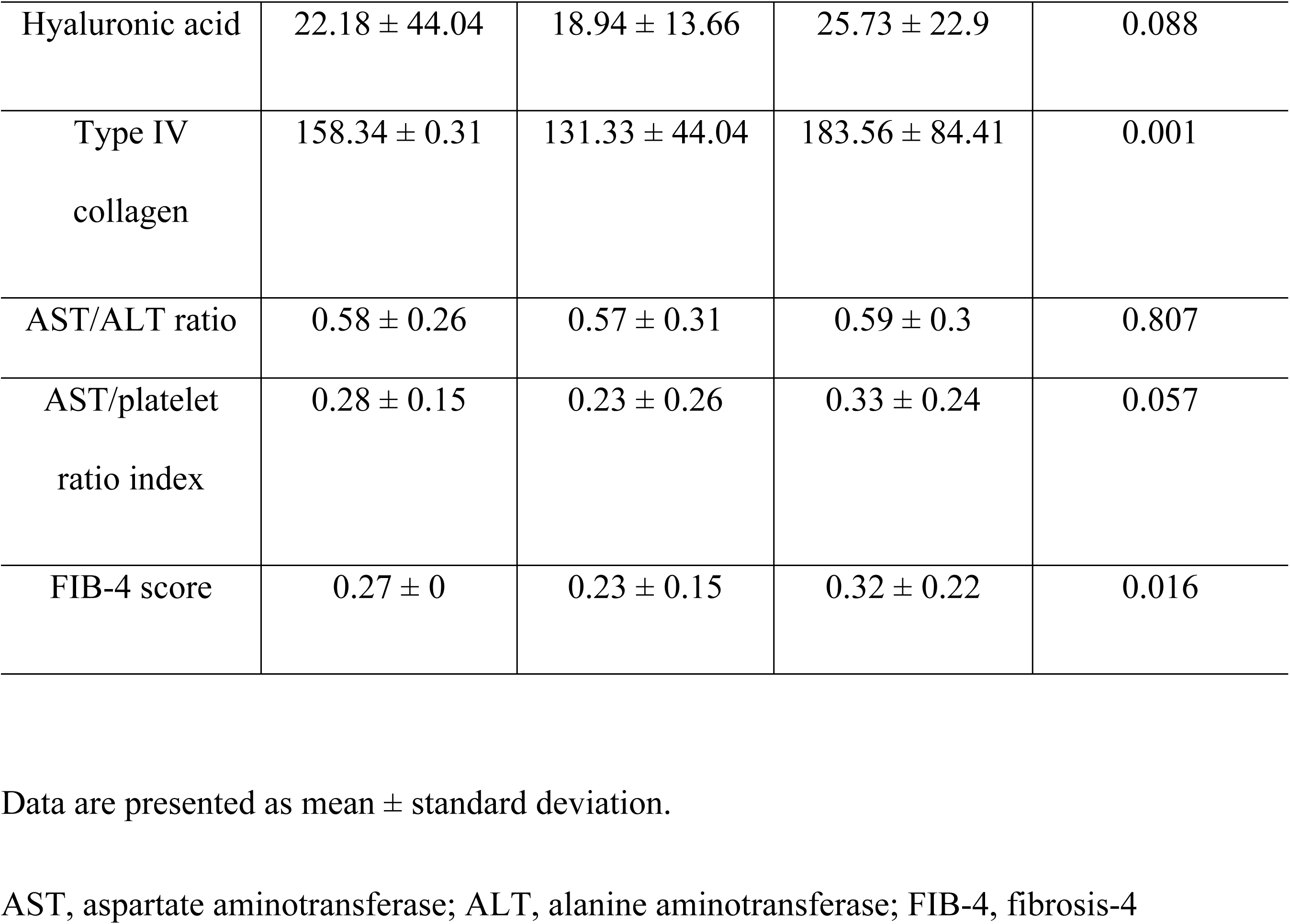
Comparison of noninvasive hepatic fibrosis markers between mild fibrosis and advanced fibrosis in obese children with nonalcoholic steatohepatitis.

In the comparison of hepatic fibrosis scores between the mild and advanced fibrosis groups, the FIB-4 score revealed statistically significant differences between the two groups (0.23 ± 0.15 vs. 0.32 ± 0.22, *p =* 0.016) (Table 2). No significant differences were observed in the AST/ALT ratio and APRI.

### Comparison of diagnostic accuracy of noninvasive hepatic fibrosis markers

The ROC curves of noninvasive serologic markers and hepatic fibrosis scoring systems in obese children with NASH are shown in Fig. 1.

**Figure 1.**
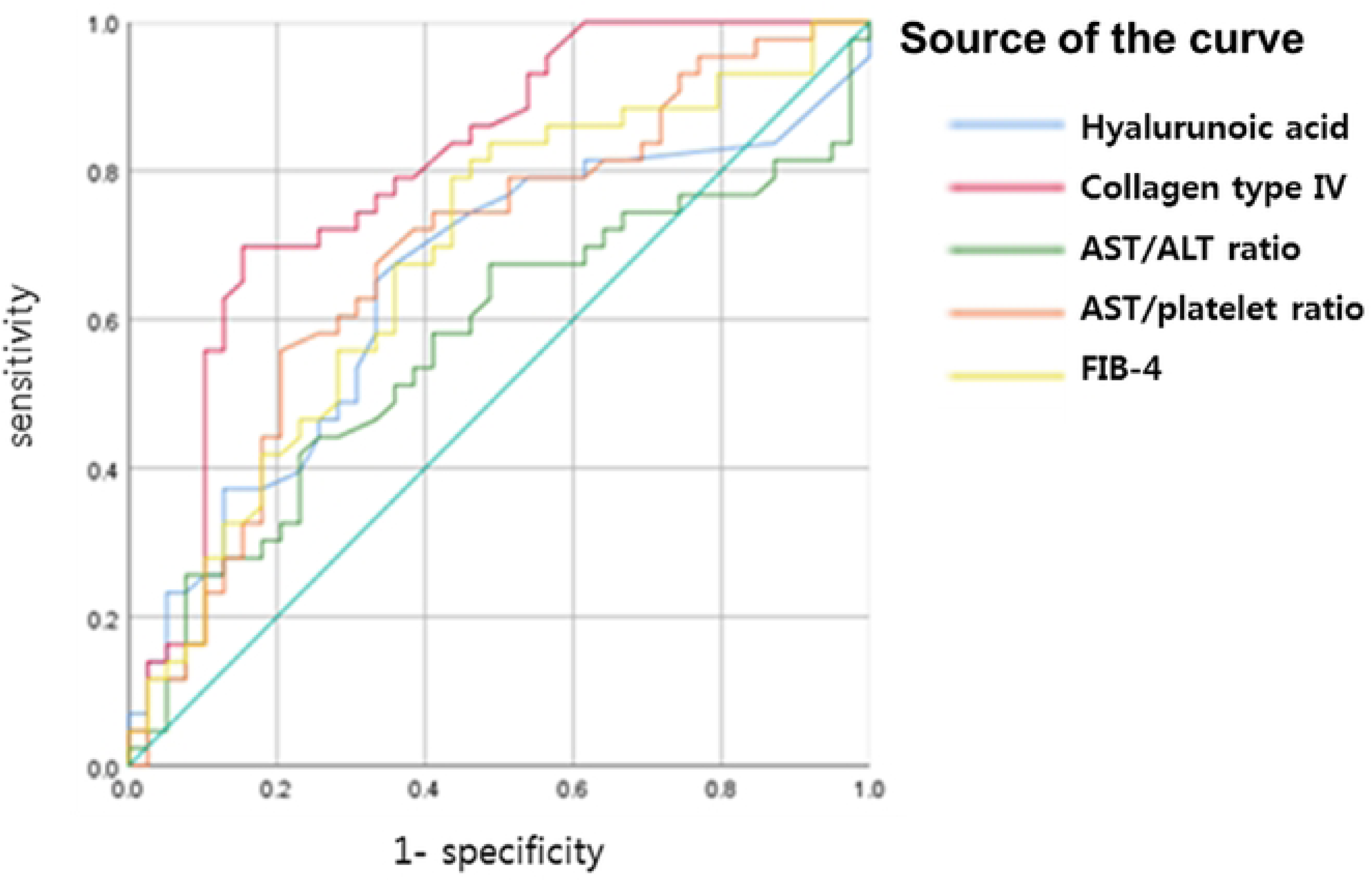
Receiver operating characteristic curves for noninvasive hepatic fibrosis scoring systems (hyaluronic acid, type 4 collagen, AST/ALT ratio, AST/platelet ratio, FIB-4) used to diagnose clinically advanced fibrosis (stage 2-4) in obese children with nonalcoholic steatohepatitis. ALT, alanine aminotransferase; AST, aspartate aminotransferase; FIB-4, fibrosis-4.

The AUROC of each biomarker and scoring system was 0.80 (95% confidence interval [CI]: 0.70-0.90) for type IV collagen at an optimal cutoff of 148 ng/mL (sensitivity 69.8%, specificity 84.6%), followed by 0.69 (95% CI: 0.57-0.83) for APRI, 0.68 (95% CI: 0.56-0.80) for FIB-4, 0.65 (95% CI: 0.53-0.77) for HA, and 0.57 (95% CI: 0.44-0.69) for the AST/ALT ratio (Fig. 1).

## Discussion

In the present study, we evaluated the diagnostic accuracy of potential serologic biomarkers related to the pathophysiology of liver fibrosis. Further, we compared the diagnostic accuracy of these significant serologic biomarkers with that of noninvasive hepatic fibrosis scoring systems according to the severity of hepatic fibrosis on liver histopathology in obese children with NASH. Our results revealed, for the first time, that type IV collagen is a useful single biomarker that can noninvasively distinguish advanced fibrosis at an optimal cutoff in pediatric patients with NASH.

Hepatic fibrosis, a dynamic process in which the contrasting processes of fibrogenesis and fibrolysis occur together, results in the deposition of collagen and extracellular matrix proteins in hepatic tissue [7]. Continued deposition of these substances results in structural changes in the liver tissue and functional disorders of the parenchyma, ultimately leading to chronic complications such as liver cirrhosis. Thus, the severity of liver fibrosis is an important prognostic factor for determining the prognosis and timing of chronic liver disease [18].

In general, liver biopsy is used to diagnose the presence and severity of hepatic fibrosis because it is the gold standard diagnostic method in both adults and children [19]. However, it has some limitations in clinical practice because of its invasiveness, sampling errors, cost, and complications, especially in pediatric patients [20]. In addition, for these reasons, it is difficult to perform repeat liver biopsies for the short-term follow-up of hepatic fibrosis [7]. Therefore, there is an increasing need to find noninvasive markers of hepatic fibrosis.

To date, a number of studies have investigated noninvasive tools for evaluating liver fibrosis in adult patients [7, 10, 21–27]. With respect to noninvasive methods of detecting hepatic fibrosis, there are two major classes: class I comprising serologic biomarkers of fibrosis and class II consisting of hepatic fibrosis scores, both of which have mainly been developed for adult patients with chronic liver diseases [21]. However, to date, validation studies on noninvasive markers that can predict the severity of liver fibrosis in children and adolescents are lacking.

For class II noninvasive hepatic fibrosis scoring systems, there have been many studies on multiparametric algorithms that have been statistically validated with respect to the detection of the presence and activity of ongoing fibrosis [21]. To date, various noninvasive hepatic fibrosis scores, including the AST/ALT ratio [12], APRI [13], PGA index [28], Forns index [29], FIB-4 [14], and NAFLD fibrosis score [30], have been developed and applied to chronic liver diseases such as hepatitis B, hepatitis C, alcoholic fatty liver disease, and NAFLD [31, 32]. These hepatic fibrosis scores, applied in adults with NAFLD, have also been validated in several previous studies [33]. However, in pediatric patients, hepatic fibrosis scores are less accurate. Moreover, not all of these scores are liver specific [26]. One disadvantage is that the scores can be affected by comorbid conditions [26]. Recently, a few validation studies, including our previous study, have been conducted in obese children with NAFLD for the noninvasive diagnosis of hepatic fibrosis using the AST/ALT ratio, APRI, PGA index, Forns index, NAFLD fibrosis score, FIB-4, and pediatric NAFLD fibrosis index [12, 25]. The results of the studies revealed that only APRI and FIB-4 score significantly distinguished advanced fibrosis (stage 2-3) from no/mild fibrosis (stage 0-1). However, the diagnostic accuracy of single biomarkers in children was not evaluated and compared [25, 34]. Even in the present study, among the noninvasive hepatic fibrosis scoring systems, FIB-4 was significantly different between the no/mild fibrosis and advanced fibrosis groups (*p* = 0.016). APRI also showed a difference between the groups, although it was not statistically significant (*p* = 0.057). Further, the diagnostic accuracy of these hepatic fibrosis scores was compared with that of potential single serologic markers in this study.

The potential serologic biomarkers for the noninvasive screening of hepatic fibrosis can be divided into direct and indirect markers. Direct markers indicate the levels of synthesis and degradation products of the extracellular matrix, such as type IV collagen and HA, whereas indirect markers such as platelet count, serum cholesterol, and transaminases do not directly reflect the metabolism of the extracellular matrix but represent hepatic function [35]. In adult studies, it has been reported that among direct markers, collagen IV has a statistically significant association with the degree of liver fibrosis [36, 22]. Yoneda M et al reported that collagen IV had AUROC of 0.828, 70% sensitivity, 81% specificity, and 86% positive predictive value [36]. In addition, hyaluronic acid (hyaluronan) has been reported as one of the sensitive tests according to several studies, which are recent investigations of cirrhosis due to nonalcoholic fatty liver disease and other etiologies [37,38,39]. Lydataki et al study reported that HA in NAFLD patients had a region of 0.97 AUROC), a sensitivity of 86% -100%, and a specificity of about 88% [37]. Oberti et al reported that HA (86%) was superior to laminin and procollagen as a result of examining the diagnostic accuracy of direct markers of liver fibrosis[38]. However, the accuracy of fibrosis biomarker in children has not been determined yet, although studies of scoring systems such as panels with the same AST to platelet ratio have been conducted [40–44].

In obese children and adolescent with NASH, there are several studies on noninvasive serological markers but most of them were about indirect markers. Moreover, no single direct serologic biomarker has been identified and applied to pediatric clinical practice thus far. Valerio NoBILI et al et al evaluated the association between HA and liver fibrosis in 100 children with NAFLD for which biopsy has been proven. In their study, values of HA> 1200 ng/mL were likely to be absence of fibrosis (F0) (7%, 95% confidence interval [CI]: 1% to 14%), values of HA> 2100 ng/mL were F2 to F4 fibrosis (89%, 95% CI: 75% to 100%) [43].

Therefore, in this study, we attempted to evaluate all possible serologic markers for hepatic fibrosis in 99 obese children diagnosed with NAFLD on liver biopsy. We evaluated all clinical and biochemical markers and compared them according to the histopathologic stages of hepatic fibrosis; however, no clinical and laboratory parameters (including age, sex, BMI, platelet count, liver enzymes, cholesterol or triglyceride, and homeostasis model assessment-estimated insulin resistance index) as indirect markers, except abdominal adiposity, were significantly different between the no/mild fibrosis and advanced fibrosis groups. However, when we compared the direct serologic markers of fibrosis based on the mechanism of hepatic fibrinogenesis, the serum levels of type IV collagen were significantly different between the two fibrosis groups, and could distinguish pediatric patients with NASH with advanced fibrosis from those with no/mild fibrosis.

In the present study, to determine the most accurate noninvasive diagnostic tools for detecting hepatic fibrosis in obese children and adolescents with NASH, we additionally compared the diagnostic accuracy and AUROCs of potential noninvasive markers, including the serum levels of HA and type IV collagen as class I serologic markers of hepatic fibrosis and FIB-4 and APRI as class II hepatic fibrosis scores, between the mild fibrosis and advanced fibrosis groups of obese children diagnosed with NAFLD. According to our findings, the AUROC of each serologic biomarker and hepatic fibrosis scoring system was 0.80 (95% CI: 0.70-0.90) for type IV collagen, which had the highest diagnostic accuracy, followed by 0.69 (95% CI: 0.57-0.83) for APRI, 0.68 (95% CI: 0.56-0.80) for FIB-4, and 0.65 (95% CI: 0.53-0.77) for HA.

Furthermore, in our study, the optimal cutoff of type IV collagen (≥ 148 ng/mL) was highly likely to distinguish advanced fibrosis from no or mild fibrosis with a sensitivity of 69.8% and a specificity of 84.6%. In recent studies on liver cirrhosis related to NAFLD or other chronic liver diseases, HA was reported to be a biomarker of hepatic fibrosis with an AUROC of 0.97, a sensitivity of 83% to 100%, and a specificity of 66% to 88% [7, 37]. In a recent study, the negative prediction (98-100%) was significantly higher than the positive prediction (61%) at a cutoff value of 60 μg/L for HA; thus, HA as a serologic marker was considered capable of excluding advanced fibrosis and cirrhosis in adult patients [37]. A study performed in children with NAFLD reported that a serum HA level of ≥ 21 μg/L indicates a high probability of developing advanced fibrosis (89%, 95% CI: 75-100%) [43]. In contrast, our study showed that the serum levels of HA did not significantly differ between the no/mild and advanced fibrosis groups, whereas the highest AUROC value was noted for serum levels of type IV collagen with significant differences between the two groups.

Our study had some limitations. As suggested in previous studies, the histopathologic findings of pediatric NASH are somewhat different from those of adult NASH [4, 5]. Thus, previous fibrosis scores developed for adults may not be completely applicable to pediatric patients. And in adult studies, there was a study comparing serum markers that could replace liver biopsy with non-serum markers such as fibroscan. Further validation studies may be needed in the future to apply noninvasive markers, not only single serologic markers but also hepatic fibrosis scores and non-serum markers, to pediatric clinical practice.

Nevertheless, our study has clinical significance because it revealed that type IV collagen is a useful single noninvasive serologic biomarker for predicting liver fibrosis in obese children with NASH. We suggested an optimal cutoff value for the serum level of type IV collagen that can distinguish an advanced hepatic fibrosis status from a no/mild fibrosis status. Furthermore, the diagnostic accuracies of the other potential noninvasive clinical and serologic markers and hepatic fibrosis scores were also evaluated in the present study. If the degree of fibrosis of NAFLD can be predicted using these noninvasive markers in clinical practice, it will be helpful in determining proper diagnostic and therapeutic strategies as soon as possible before hepatic fibrosis progresses. Therefore, in the future, studies on noninvasive markers of liver fibrosis need to be conducted, especially in children.

## Conclusion

In the current study, Type IV collagen as a single noninvasive serologic biomarker for hepatic fibrosis and FIB-4 as a hepatic fibrosis score are beneficial in predicting advanced hepatic fibrosis, and in determining proper diagnosis and treatment strategies before fibrosis progresses in obese children with NASH.

## Data Availability

I have attached the data as a Supporting information file. (File name : data_PLOS ONE)

## Abbreviations

NAFLD: Nonalcoholic fatty liver disease
NASH: nonalcoholic steatohepatitis
BMI: Body Mass Index
AC: Abdominal Circumference
AST: ASpartate aminoTransferase
ALT: ALanine aminotransferase
HA: hyaluronic acid
APRI: AST-to-platelet ratio index
FIB-4: Fibrosis-4 index
ROC: Receiver-Operating Characteristic
AUROC: Area Under the ROC curve
CI: Confidence Interval

## Declarations

### Availability of Data and Material

The datasets used and/or analyzed in the current study are available from the corresponding author on reasonable request.

### Author contributions

Jung Yeon Joo: conducting the study, drafting the manuscript In Hyuk Yoo: collecting and interpreting data

Hye Ran Yang: designing the study, interpreting data, drafting the manuscript

## References

1. Oh K, Jang MJ, Lee NY, Moon JS, Lee CG, Yoo MH et al. Prevalence and trends in obesity among Korean children and adolescents in 1997 and 2005. Korean J Pediatr 2008;51:950–5.

2. Bellentani S, Scaglioni F, Marino M, Bedogni G. Epidemiology of non-alcoholic fatty liver disease. Dig Dis 2010;28:155–61.

3. Barshop NJ, Sirlin CB, Schwimmer JB, Lavine JE. Review article: epidemiology, pathogenesis and potential treatments of paediatric non-alcoholic fatty liver disease. Aliment Pharmacol Ther 2008;28:13–24.

4. Miriam B. Vos, Stephanie H. Abrams, Sarah E. Barlow, Sonia C., Stephen R. Daniels, Rohit Kohli et al. NASPGHAN Clinical Practice Guideline for the Diagnosis and Treatment of Nonalcoholic Fatty Liver Disease in Children: Recommendations from the Expert Committee on NAFLD (ECON) and the North American Society of Pediatric Gastroenterology, Hepatology and Nutrition (NASPGHAN). J Pediatr Gastroenterol Nutr. 2017 Feb;64(2):319–334.

5. Powell EE, Cooksley WG, Hanson R, Searle J, Halliday J W, Powell L W. The natural history of nonalcoholic steatohepatitis: a follow-up study of forty-two patients for up to 21 years. Hepatology 1990; 11: 74–80.

6. Molleston JP, White F, Teckman J, Fitzgerald JF. Obese children with steatohepatitis can develop cirrhosis in childhood. Am J Gastroenterol 2002; 97: 2460–2.

7. Don C. Rockey, D. Montgomery Bissell. Noninvasive Measures of Liver Fibrosis. Hepatology 2006 Feb;43(2 Suppl 1):S113–20.

8. Straub BK, Schirmacher P. Pathology and biopsy assessment of non-alcoholic fatty liver disease. Dig Dis 2010; 28: 197–202.

9. Schwimmer JB, Behling C, Newbury R, Deutsch R, Nievergelt C, Schork NJ, Lavine JE. Histopathology of pediatric nonalcoholic fatty liver disease. Hepatology 2005;42(3):641–9.

10. Guha IN, Parkes J, Roderick PR, Harris S, Rosenberg WM. Non-invasive markers associated with liver fibrosis in nonalcoholic fatty liver disease. Gut 2006; 55: 1650–60.

11. Kim JH, Yun S, Hwang SS, Shim JO, Chae HW, Lee YJ, et al. The 2017 Korean National Growth Charts for children and adolescents: development, improvement, and prospects. Korean J Pediatr 2018;61:135–49.

12. Iacobellis A, Marcellini M, Andriulli A, Perri F, Leandro G, Devito R et al. Non invasive evaluation of liver fibrosis in paediatric patients with nonalcoholic steatohepatitis. World J Gastroenterol 2006;12:7821–5.

13. Loaeza-del-Castillo A, Paz-Pineda F, Oviedo-Cárdenas E, Sánchez-Avila F, Vargas- Vorácková F. AST to platelet ratio index (APRI) for the noninvasive evaluation of liver fibrosis. Ann Hepatol 2008;7:350–7.

14. Vallet-Pichard A, Mallet V, Nalpas B, Verkarre V, Nalpas A, Dhalluin-Venier V et al. FIB- 4: an inexpensive and accurate marker of fibrosis in HCV infection. comparison with liver biopsy and fibrotest. Hepatology 2007;46: 32–6.

15. Poynard T, Aubert A, Bedossa P, Abella A, Naveau S, Paraf F et al. A simple biological index for detection of alcoholic liver disease in drinkers. Gastroenterology 1991;100:1397–402.

16. Kim JM. Pathologic diagnosis of hepatic fibrosis. Clinical and Molecular Hepatology 2006;4s:69–74.

17. Strauss S, Gavish E, Gottlieb P, Katsnelson L Interobserver and intraobserver variability in the sonographic assessment of fatty liver. AJR Am J Roentgenol 2007;189:W320–3.

18. Nobili V, Pinzani M. Paediatric non-alcoholic fatty liver disease. Gut 2010;59:561–4.

19. Kleiner DE, Brunt EM, Van Natta M, Behling C, Contos MJ, Cummings OW et al. Design and validation of a histological scoring system for nonalcoholic fatty liver disease. Hepatology 2005;41:1313–21.

20. Nobili V, Comparcola D, Sartorelli MR, Natali G, Monti L, Falappa P, Marcellini M. Blind and ultrasound-guided percutaneous liver biopsy in children. Pediatr Radiol 2003;33:772–5.

21. Gressner AM, Gao CF, Gressner OA. Non-invasive biomarkers for monitoring the fibrogenic process in liver: A short survey World J Gastroenterol 2009;28;15:2433–40.

22. Lesmana CRA, Hasan I, Bundihusodo U, Gani RA, Krisnuhoni E, Akbar N, Lesmana LA. Diagnostic value of a group of biochemical markers of liver fibrosis in patients with non-alcoholic steatohepatitis. J Dig Dis 2009;10;201–6

23. Alkhouri N, Carter-Kent C, Lopez R, Rosenberg WM, Pinzani M, Bedogni G et al. A combination of the pediatric NAFLD fibrosis index and enhanced liver fibrosis test identifies children with fibrosis. Clin Gastroenterol Hepatol 2011;9:150–5.

24. Alkhouri N, De Vito R, Alisi A, Yerian L, Lopez R, Feldstein AE et al. Development and validation of a new histological score for pediatric non-alcoholic fatty liver disease. J Hepatol. 2012;57:1312–8.

25. Yang HR, Kim HR, Kim MJ, Ko JS, Seo JK. Noninvasive parameters and hepatic fibrosis scores in children with nonalcoholic fatty liver disease. World J Gastroenterol 2012;18:1525–30.

26. Castera L, Friedrich-Rust M, Loomba R. Noninvasive Assessment of Liver Disease in Patients With Nonalcoholic Fatty Liver Disease.. Gastroenterology 2019;156:1264–81

27. Enomoto H, Bando Y, Nakamura H, Nishiguchi S, Koga M. Liver fibrosis markers of nonalcoholic steatohepatitis. World J Gastroenterol 2015;28;21:7427–35.

28. Teare JP, Greenfield SM, Thompson R.P.H., Simpson J, Sherman D, Bray G et al. Comparison of serum procollagen III peptide concentrations and PGA index for assessment of hepatic fibrosis. Lancet 1993; 342: 895–8

29. Forns X, Ampurdanès S, Llovet JM, Aponte J, Quintó L, Martınez-Baucer E et al. Identification of chronic hepatitis C patients without hepatic fibrosis by a simple predictive model. Hepatology 2002;36 986–92.

30. Angulo P, Hui JM, Marchesini G, Bugianesi E, George J, Farrell GC et al. The NAFLD fibrosis score: a noninvasive system that identifies liver fibrosis in patients with NAFLD. Hepatology 2007;45:846–54.

31. Poynard T, Morra R, Ingiliz P, Imbert-Bismut F, Thabut D, Messous D, et al. Biomarkers of liver fibrosis. Adv Clin Chem 2008;46: 131–60.

32. Adams LA. Biomarkers of liver fibrosis. J Gastroenterol Hepatol 2011;26:802–9

33. Maurice J, Manousou P. Non-alcoholic fatty liver disease. Clin Med 2018 Vol 18, No 3: 245–50

34. Ko JS, Yoon JM, Yang HR, Myung JK, Kim HY, Kang GH et al Clinical and Histological Features of Nonalcoholic Fatty Liver Disease in Children. Dig Dis Sci 2009;54:2225–30.

35. Carey E, Carey WD. Noninvasive tests for liver disease, fibrosis, and cirrhosis: Is liver biopsy obsolete? Cleveland Clinic Journal of Medicine August 2010;77:519–27

36. Yoneda M, Mawatari H, Fujita K Yonemitsu K, Kato S, Takahashi H, et al. Type IV collagen 7s domain is an independent clinical marker of the severity of fibrosis in patients with nonalcoholic steatohepatitis before the cirrhotic stage. J Gastroenterol 2007; 42: 375–81.

37. Lydatakis H, Hager IP, Kostadelou E, Mpousmpoulas S, Pappas S, Diamantis I et al. Non-invasive markers to predict the liver fibrosis in non-alcoholic fatty liver disease. Liver Int. 2006;26:864–71.

38. Oberti F, Valsesia E, Pilette C, Rousselet MC, Bedossa P, Aube C, et al. Noninvasive diagnosis of hepatic fibrosis or cirrhosis. Gastroenterology 1997;113:1609–16.

39. Suzuki A, Angulo P, Lymp J Li D, Satomura S, Lindor K. Hyaluronic acid, an accurate serum marker for severe hepatic fibrosis in patients with non-alcoholic fatty liver disease. Liver Int 2005; 25: 779–86.

40. Kim E, Kang Y, Hahn S, Lee MJ, Park YN, Koh H. The efficacy of aspartate aminotransferase-toplatelet ratio index for assessing hepatic fibrosis in childhood nonalcoholic steatohepatitis for medical practice. Korean journal of pediatrics. 2013;56:19–25.

41. Flores-Calderon J, Moran-Villota S, Ramon-Garcia G, González-Romano B, Bojórquez- Ramos BC, Cerdán-Silva L et al. Non-invasive markers of liver fibrosis in chronic liver disease in a group of Mexican children. A multicenter study. Annals of hepatology. 2012;11:364–8.

42. Kaneda H, Hashimoto E, Yatsuji S, Tokushige K, Shiratori K. Hyaluronic acid levels can predict severe fibrosis and platelet counts can predict cirrhosis in patients with nonalcoholic fatty liver disease. Journal of gastroenterology and hepatology. 2006;21:1459–65.

43. Nobili V, Alisi A, Torre G, Vito RD, Pietrobattista A, Morino G et al. Hyaluronic acid predicts hepatic fibrosis in children with nonalcoholic fatty liver disease. Transl Res. 2010;156:229–34.

